# Generation of realistic virtual adult populations using a model-based copula approach

**DOI:** 10.1101/2024.02.22.24303086

**Authors:** Yuchen Guo, Tingjie Guo, Catherijne A.J. Knibbe, Laura B. Zwep, J.G. Coen van Hasselt

## Abstract

Incorporating realistic sets of patient-associated covariates, i.e., virtual populations, in pharmacometric simulation workflows is essential to obtain realistic model predictions. Current covariate simulation strategies often omit or simplify dependency structures between covariates. Copula models are multivariate distribution functions suitable to capture dependency structures between covariates with improved performance compared to standard approaches. We aimed to develop and evaluate a copula model for generation of adult virtual populations for 12 patient-associated covariates commonly used in pharmacometric simulations, using the publicly available NHANES database, including sex, race-ethnicity, body weight, albumin, and several biochemical variables related to organ function. A multivariate (vine) copula was constructed from bivariate relationships in a stepwise fashion. Covariate distributions were well captured for the overall and subgroup populations. Based on the developed copula model, a web application was developed. The developed copula model and associated web application can be used to generate realistic adult virtual populations, ultimately to support model-based clinical trial design or dose optimization strategies.

## 1. Introduction

In pharmacometric modeling, patients’ covariates are usually identified as a source of variability between individuals that impacts pharmacokinetics and pharmacodynamics [1]. Generation of virtual populations (VPs), i.e., realistic sets of patient characteristics or covariates, is essential to ensure that realistic responses are produced in pharmacometric simulations, eventually providing valuable information to support *in silico* clinical trials and optimization of dosing strategies.

Realistic VPs should reflect both the marginal distribution and dependency structure observed between covariate variables of interest. In statistics, a marginal distribution describes the probability distribution of one separate variable, and a dependence structure reveals the relationships or dependent patterns between variables in a dataset. For instance, within a real-world dataset focusing on the elderly population, age, as a covariate, may exhibit a t-distributed margin, with a certain mean and standard deviation; meanwhile, it could be negatively correlated with renal function biomarkers. Misspecification of the margins or dependency structures, i.e., in comparison with those actually observed, may impact the quality of subsequent patient responses obtained in pharmacometric simulations.

VPs can be generated using several approaches, which are either data-driven or distribution-driven. Data-driven methodologies such as the bootstrap or conditional distribution modeling [4, 5] utilize an actual dataset of patient characteristics to sample from. Requesting such data, however, is sometimes not possible due to patient privacy regulations. Distribution-based approaches characterize the distribution of the marginals of covariates of interest but may not always capture their dependency structure. For example, series of univariate distributions can be used to describe the marginals yet ignore interdependencies between covariates. Multivariate normal distributions [3] do consider the dependency but assume that variables are normally distributed, which may not always hold. Finally, machine learning algorithms [4–6] have been proposed, but these models are usually based on complex frameworks and often lack interpretability of underlying dependencies.

Copula models are statistical models which capture the dependence structure between random variables independently from the description of the marginals [7]. A rich variety of copula models is available to be selected to estimate diverse dependent patterns in data [8]. Using a transformation of any marginal distribution to a uniform distribution, the dependence structure can be separated from the marginal structure. An extension of the copula, the vine copula, addresses the difficulty of calculating multivariate joint distributions by using conditional dependence and bivariate building blocks [9]. We have previously proposed the copula as a relevant key strategy for VP generation, demonstrating favorable performance in simulating realistic VPs compared to standard approaches, while their distribution-based nature facilitates sharing of covariate data within the community [10].

Here, we present a copula model for simulation of adult virtual populations. We first developed a copula model for 12 covariates of relevance for pharmacometric models using data from adult individuals present in the NHANES database [11]. Then we evaluated the performance of the copula in simulating the overall and subgroup populations. Finally, a web application was designed for the copula model developed to facilitate generation of adult VPs.

## 2. Methods

### 2.1 Data

We used the public database from National Health and Nutrition Examination Survey (NHANES), an initiative that collects data on non-institutionalized individuals in the U.S., including laboratory measurements, physical screening, and surveys; data are released to the public every two years[11]. We combined the NHANES data for 2009∼2010, 2011∼2012, 2013∼2014, 2015∼2016, and 2017∼2018 releases based on their accessibility and consistency in laboratory methods. Differences in laboratory, instruments, and methods across releases were considered by implementing the adjustment equations provided by NHANES.

We focused on the adult population aged 18-80 years, with 27,008 subjects in total. Common covariates of interest for population pharmacokinetic models were selected: sex, race-ethnicity, age, height, body weight, fat mass (Fat), serum creatinine (SCR), alanine aminotransferase (ALT), aspartate aminotransferase (AST), alkaline phosphatase (ALP), albumin and total bilirubin (BR) [12–16]. We acknowledge the sensitivity regarding the use of race-ethnicity as a medical indicator. Its inclusion in our study is focused on subgroup analysis when relevant, and not intended to perpetuate stereotypes or contribute to health disparities.

Table 1 provides the summary statistics of covariates in the model development dataset. Of note, over 50% of fat mass data in the observed dataset were missing. Half of the missing data were due to not meeting the inclusion criteria of age (< 60 years old) during the examination, while another half were due to the examination not being conducted in the 2009∼2010 release. Since the copula approach allows to impute missing values, imputation analysis regarding fat mass data (**supplementary material**) was performed to provide more insights into the reliability of simulated fat mass data for people aged ≥ 60 years, and the result showed no significant bias in the simulated fat mass for people above 60 years old (**Figure S4**).

**Table 1.** Summary statistics of covariates in dataset combined from National Health and Nutrition Examination Surveys 2009∼2010, 2011∼2012, 2013∼2014, 2015∼2016 and 2017∼2018. The total number of individuals was n = 27008.

| VARIABLE NAME | VARIABLE DESCRIPTION | PERCENTAGE (%) | ACTUAL N (% MISSING) | MEAN ± SD [RANGE] |
| --- | --- | --- | --- | --- |
| Sex | **Gender** | - | 27008 (0%) | - |
|  | Male | 48.5 | 13104 | - |
|  | Female | 51.5 | 13904 | - |
| Race-Ethnicity | **Race-Ethnicity** | - | 27008 (0%) | - |
|  | Hispanic* | 26.6 | 7176 | - |
|  | White | 36.9 | 9978 | - |
|  | African American | 22.8 | 6166 | - |
|  | Asian | 10.6 | 2859 | - |
|  | Other Race | 3.1 | 829 | - |
| Age | Age (year) | | 27008 (0%) | 46.05±17.21 [18~79] |
| Weight | Body weight (kg) | | 26746 (1.0%) | 82.02±22.17 [32.3~242.6] |
| Height | Standing height (cm) | | 26757 (1.0%) | 167.16±10.09 [123.3~204.5] |
| Fat | Total body fat (kg) | | 11826 (56.2%) | 27.11±11.93 [4.9~102.3] |
| SCR | Serum creatinine (mg/dL) | | 25313 (6.3%) | 0.88±0.45 [0.16~17.41] |
| ALT | Alanine Aminotransferase (ALT, U/L) | | 25307 (6.3%) | 25.57±20.42 [5~1363] |
| AST | Aspartate Aminotransferase (AST, U/L) | | 25288 (6.4%) | 25.93±17.13 [7~882] |
| ALP | Alkaline Phosphatase (ALP, U/L) | | 25310 (6.3%) | 69.34±24.59 [7~907] |
| Albumin | Albumin (g/dL) | | 25315 (6.3%) | 4.28±0.35 [2~5.6] |
| BR | Total bilirubin (mg/dL) | | 25298 (6.3%) | 0.62±0.31 [0~7.3] |

\* Mexican American and other Hispanic in NHANES were recorded as Hispanic in our real-world dataset. Other race-ethnicity groups remained unchanged.

### 2.2 Vine copula model development

A vine copula was fitted to the NHANES data. First, to avoid producing covariates of negative values in VPs, biochemical measurement data were log-transformed. As copulas are joint distribution functions with uniform margins, data were then transformed into uniform distributions using the probability integral function [17] based on kernel density estimation. Parametric bivariate copulas, such as Gaussian, Clayton and Frank, served as building blocks of candidate vine copula models. The vine copula model was constructed by first selecting a tree structure, which defines the pairs of covariates and copulas to be estimated. The tree structure was determined using the maximum spanning tree algorithm, which selects a tree structure that maximizes the sum of correlations [18]. Next, a bivariate copula model was selected and estimated for each copula pair in the tree. To incorporate the covariate ‘race-ethnicity’ in the copula and optimize the model, we treated race-ethnicity as an ordered categorical variable and tested copulas with all order combinations. The model was selected by minimizing the Akaike information criterion (AIC).

### 2.3 Model Evaluation

Model evaluation was conducted through a simulation-based strategy: performing 100 simulations of the original dataset and comparing the metrics between the real-world population and VPs that were back transformed to their original scales. To assess the model performance on the marginal distributions, we evaluated observed and simulated populations by comparing the frequency of each category for categorical covariates, and for continuous covariates, comparing the marginal metrics, mean, standard deviation (SD), and percentiles (5^th^, 50^th^ and 95^th^), denoted by *M*, between observed and simulated data in terms of relative error (*RE*) (**Eq.1**).

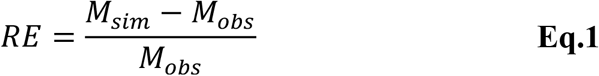

where *M*_*sim*_ and *M*_*obs*_ represent the metrics for simulated population and observed population, respectively.

To assess the performance of the model on capturing the dependency structure, pairwise correlation coefficients were compared between observed and simulated datasets. Since data sharing the same correlation could display various shapes of the dependence, a two-dimensional metric was developed to quantify the overlap of the density contours in observed and simulated data. For each pair combination of covariates, 95^th^ percentile density contours were calculated for observed and simulated populations. The overlap metric was computed as the Jaccard index: the ratio between the intersection area and union area (**Figure S1**). Higher overlap indicated a better description of dependence relations. We systematically evaluated the performance of the model from the following aspects:

(1) Overall performance: the NHANES copula (full copula) was developed based on the whole set of participants of NHANES that represents a general population. Simulated populations and the real-world population were then compared.

(2) Subgroup performance: populations of interest in clinical trials and cohort studies typically comprises individuals with certain race-ethnicity or sex. To be able to create realistic VP of interest, it is important to determine whether the full copula could capture the characteristics of subgroup populations. Predictive performance of the full copula for subsets of VPs was assessed with a particular interest in the race-ethnicity and sex subgroups. For comparison, two series of subgroup copulas were also constructed using data specific of each subgroup population :1) Hispanic copula, White copula, African American copula, Asian copula, Other race copula, 2) male copula, female copula.

Virtual subgroup populations were obtained in two ways: by simulating from the full copula model and filtering out the irrelevant individuals, and by directly simulating from the subgroup copula. The performance of full copula was compared with that of subgroup copula to provide an understanding of whether the full copula was sufficient for generating subsets of VPs.

### 2.4 Shiny Application Development

To provide a convenient and user-friendly tool, an interactive web application that could output VPs was developed using the NHANES copula. Adult virtual populations can be generated from the application.

### 2.5 Software

The analysis was performed in R 4.1.2. Processing of NHANES data was conducted with *survey* package. Kernel density estimation of marginal distributions was performed with *kde1d* package. Development of NHANES copula was implemented with *rvinecopulib* package. The overlap metric was calculated using *ks* and *sf* packages. R shiny application was developed using *shiny* package. Visualizations of this study were generated with *ggplot2* package. All scripts are available on https://github.com/vanhasseltlab/NHANES_copula.

## 3. Results

### 3.1 Vine copula of NHANES data

Logarithmic and uniform transformed data were fitted to estimate the underlying dependency structure with a vine copula. Instead of displaying the whole tree structure, we only showed the first tree since the first layer dependence captured the strongest correlations while trees of higher levels describe the conditional dependence, and are less influential on the overall fit than the first tree[19]. Sex was located at the center of the first tree structure, as it showed relatively strong dependence relationships with height, logBR, logALT, logAlbumin, and logSCR (**Figure 1A**). The density contours of covariate pairs in the real-world population displayed various patterns, and the VP were found to overlap the real-world population in selected covariate pairs well (**Figure 1B**).

**Figure 1.**
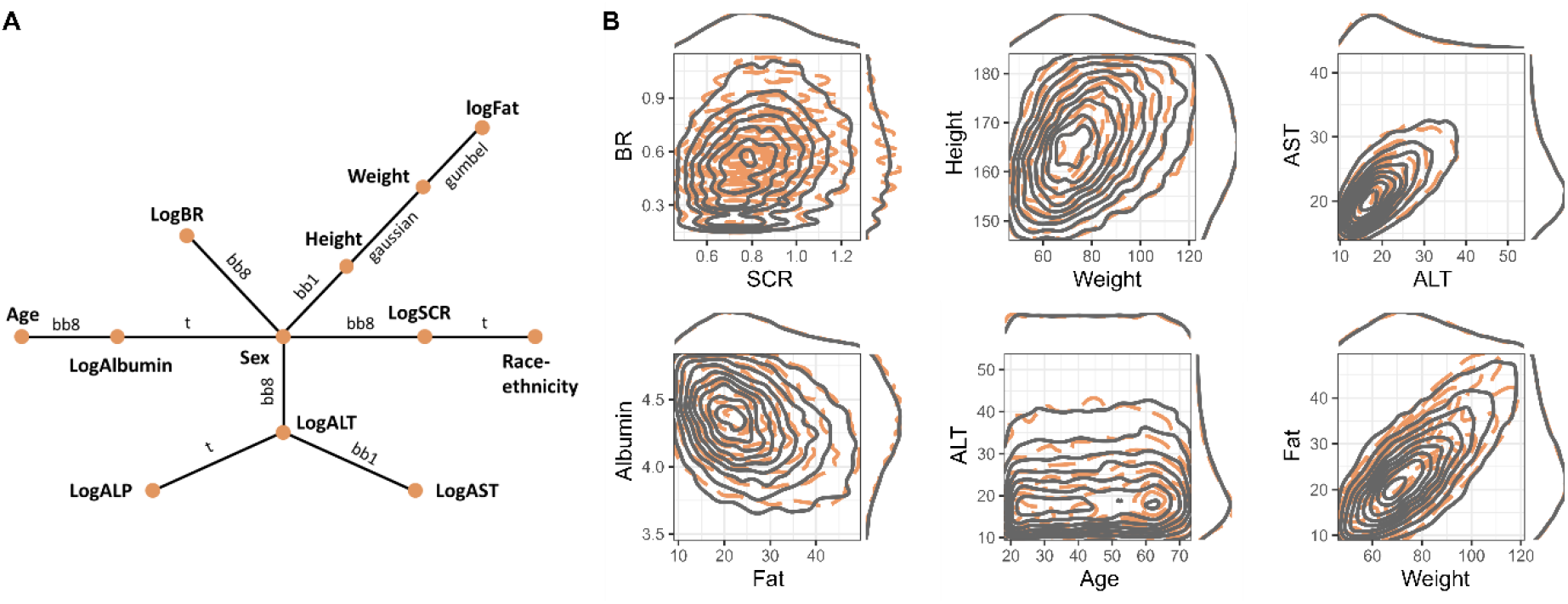
Graphical representation of dependence structure estimated by NHANES copula and bivariate densities of observed and simulated covariates. **A**. The first tree structure of NHANES copula with 12 nodes, and 11 edges. Each node represents one covariate variable, and each edge is associated with a specific bivariate copula model. 12 covariates were shown: sex, race-ethnicity, age, height, body weight, fat mass (Fat), serum creatinine (SCR), alanine aminotransferase (ALT), aspartate aminotransferase (AST), alkaline phosphatase (ALP), albumin and total bilirubin (BR). Log indicated that the covariate was log-transformed in estimation. **B**. Set of selected covariate pairs with the density contours of the observed population (orange dashed line) and the simulated population from the NHAENS copula (gray solid lines). Marginal densities were displayed on the top and right sides of each plot.

### 3.2 Overall performance

The overall simulation performance of the developed copula model was evaluated for the entire population, without specifying any subgroups. For categorical covariates, (i.e., race-ethnicity and sex), the frequency of each category in the virtual population aligned with that of the real-world population (**Figure 2A**). For continuous covariates, density curves of each individual covariate in the simulation dataset well tracked observed ones (**Figure 2B**); mean, standard deviation and percentiles of VP agreed with those of the observed population, with relative errors within ±0.10 (**Figure 2C**). For percentiles and mean metrics, coefficient of variation across simulations were all within 0.007, and those of standard deviation were within 0.09.

**Figure 2.**
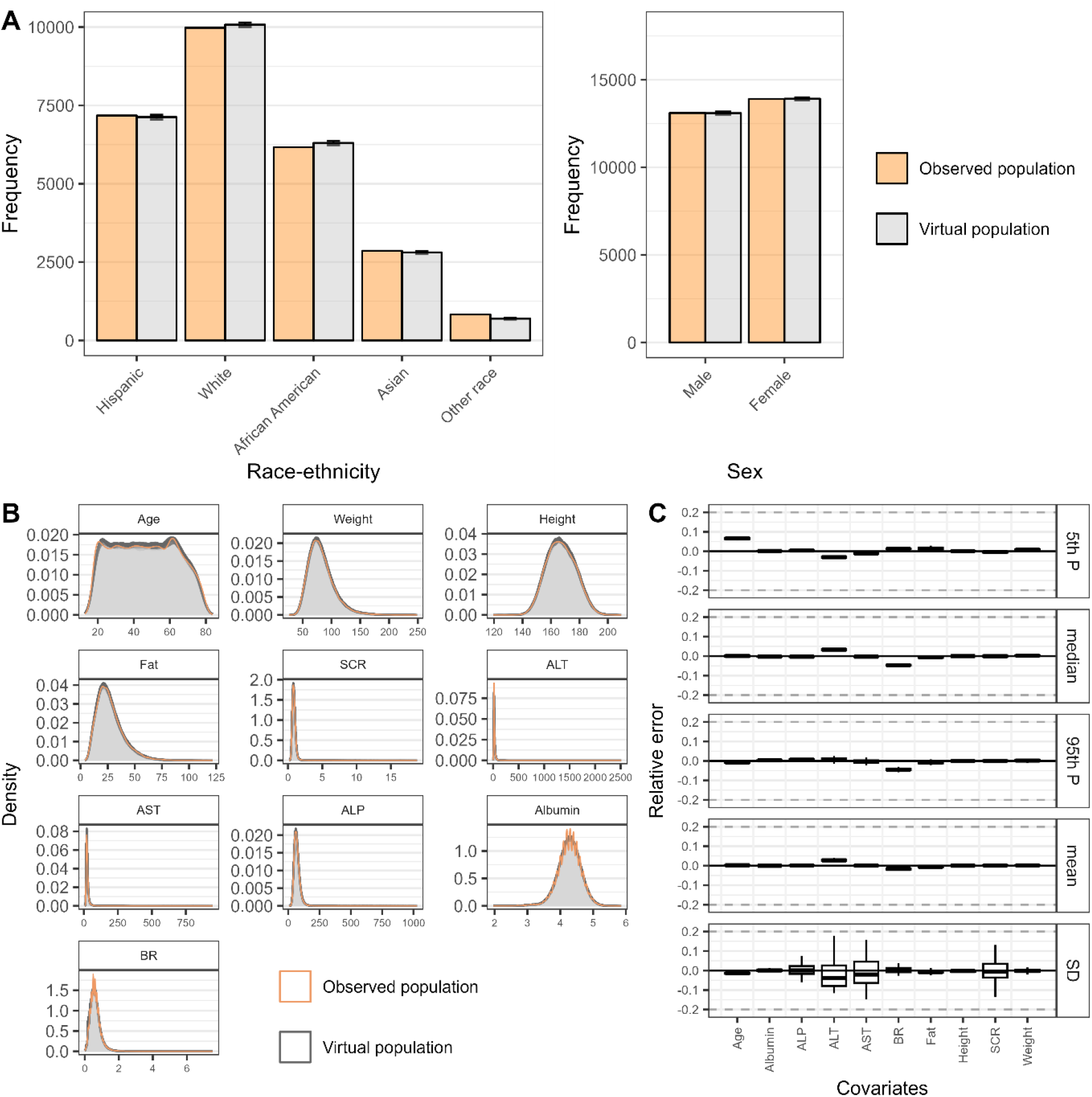
Marginal characteristics of the covariates in observed population and virtual populations simulated from NHANES copula. **A**. Frequency of each category in discrete covariates, race-ethnicity and sex, in the real-world population (orange columns) and virtual populations (grey columns). **B**. Density curves of each continuous covariate variable in the real-world population (orange line) and virtual populations (gray lines). **C**. Relative error of marginal metrics (percentiles, mean, and standard deviation) of continuous covariates as compared to the statistics of the real-world population. Virtual population was simulated 100 times. Error bars indicated the standard deviation of 100 simulations. Gray dashed lines indicate ±20% relative error.

The simulated correlations from the copula model were very similar to observed correlations for most pair combinations of covariates, with 0.023 median error (**Figure 3A**). Covariate pairs associated with the largest error of correlation were height-SCR (0.105) and SCR-albumin (0.102). The median overlap was 92.0% across all covariate pairs and simulations, and the model achieved over 85% overlap in 96% (43/45) covariate pairs, indicating a good capture of dependency structure (**Figure 3B**). The only two covariate pairs that did not reach 85% were ALT-BR and weight-fat, with 81.4% and 70.5% overlap percentages.

**Figure 3.**
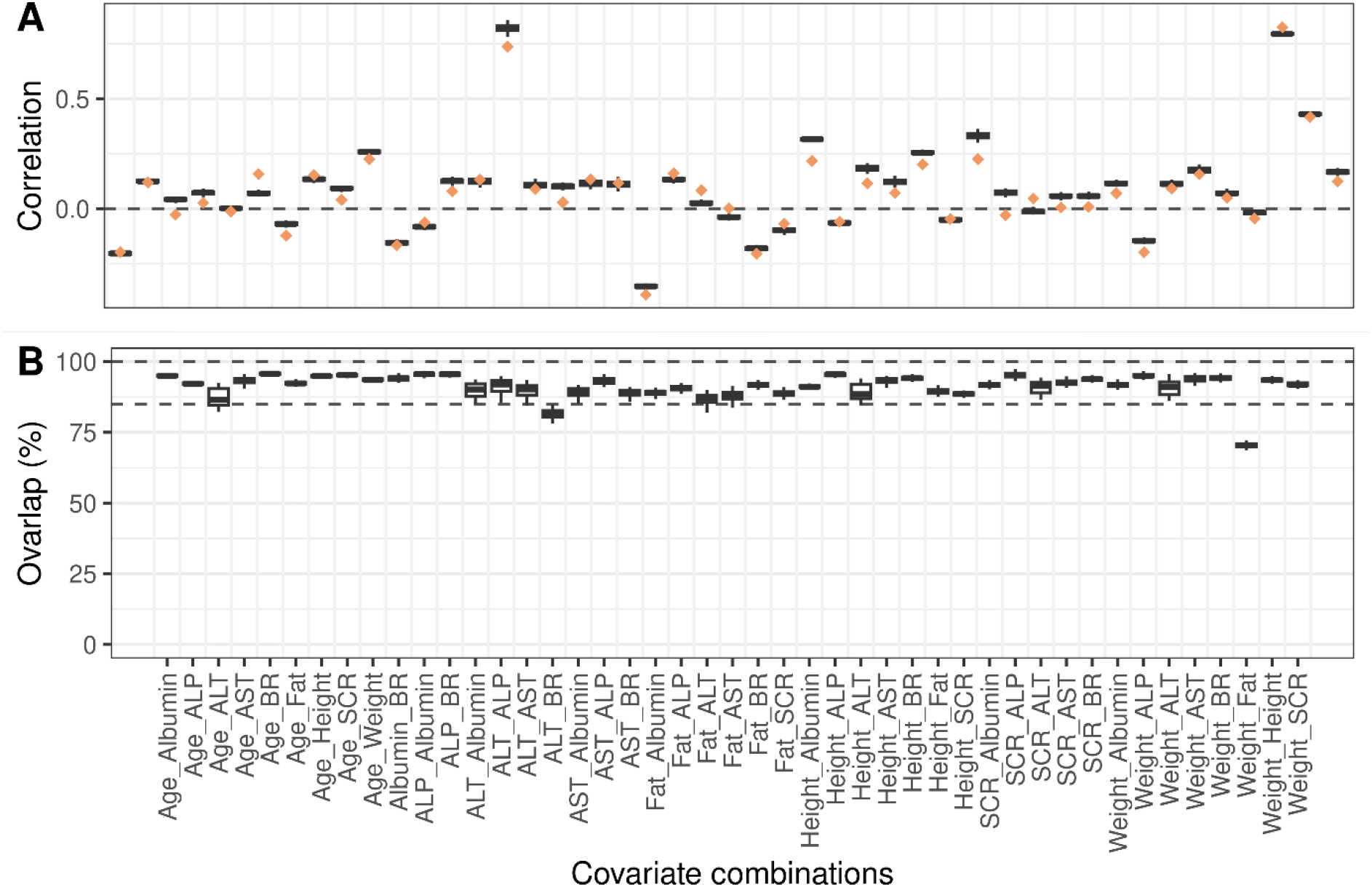
Dependency metrics of covariate pairs in observed population and virtual populations simulated from NHANES copula. **A**. Correlations of each covariate pair in real-world population (orange diamond) and virtual populations (black box). Gray dashed line represents no correlation between covariate pairs. **B**. Overlap metric of 95^th^ density contours of virtual population relative to observed population. Virtual population was simulated 100 times. Error bars indicated the standard deviation of 100 simulations. Gray dashed lines indicate 100% and 85% overlap percentages.

The full copula model reproduced the marginal properties as well as the dependence relations of covariates of input population data. Variability across simulations tended to be small for all metrics except for standard deviation, showing the robustness of the copula model.

### 3.3 Subgroup performance

To gain further insights into the usefulness of the full copula for simulating subgroups of the total population, we conducted two separate investigations on the performance of full copula for VP simulation in race-ethnicity and sex subgroups.

#### 3.3.1 Race-ethnicity subgroup analysis

The full copula was able to approximate the marginal characteristics of the observed population in Hispanic, White, and African American subgroups, with median relative errors of marginal metrics across covariates within [-0.19, 0.28] (**Figure S2**). For Asian and Other race VP populations, median relative errors were in the ranges [-0.21, 0.41] and [-0.68, 0.20], respectively. For comparison, subgroup copulas for Hispanic, White, African American and Asian populations showed good performances in terms of the marginal metrics, with the median relative errors of all covariates in [-0.15, 0.38]. However, the relative errors were larger for Other race subgroup copula, with a range of [-0.14, 0.75]. Full copula and subgroup copulas showed comparable performance in capturing the marginal attributes of Hispanic, White, African American and Other race subgroups, however, subgroup copula showed superior performance in Asian population.

The full copula model achieved 84.6%, 88.6%, 87.8%, 74.0% and 80.1% median overlap percentages for Hispanic, White, African American, Asian and Other race populations, while subgroup copulas reached 89.3%, 89.4%, 88.1%, 87.8% and 85.0% (**Figure 4A**), respectively. Subgroup copulas outperformed the full copula in simulating the dependence structure of covariates in Asian and Other race subgroups, but showed similar performance in the rest of race-ethnicity subgroups.

**Figure 4.**
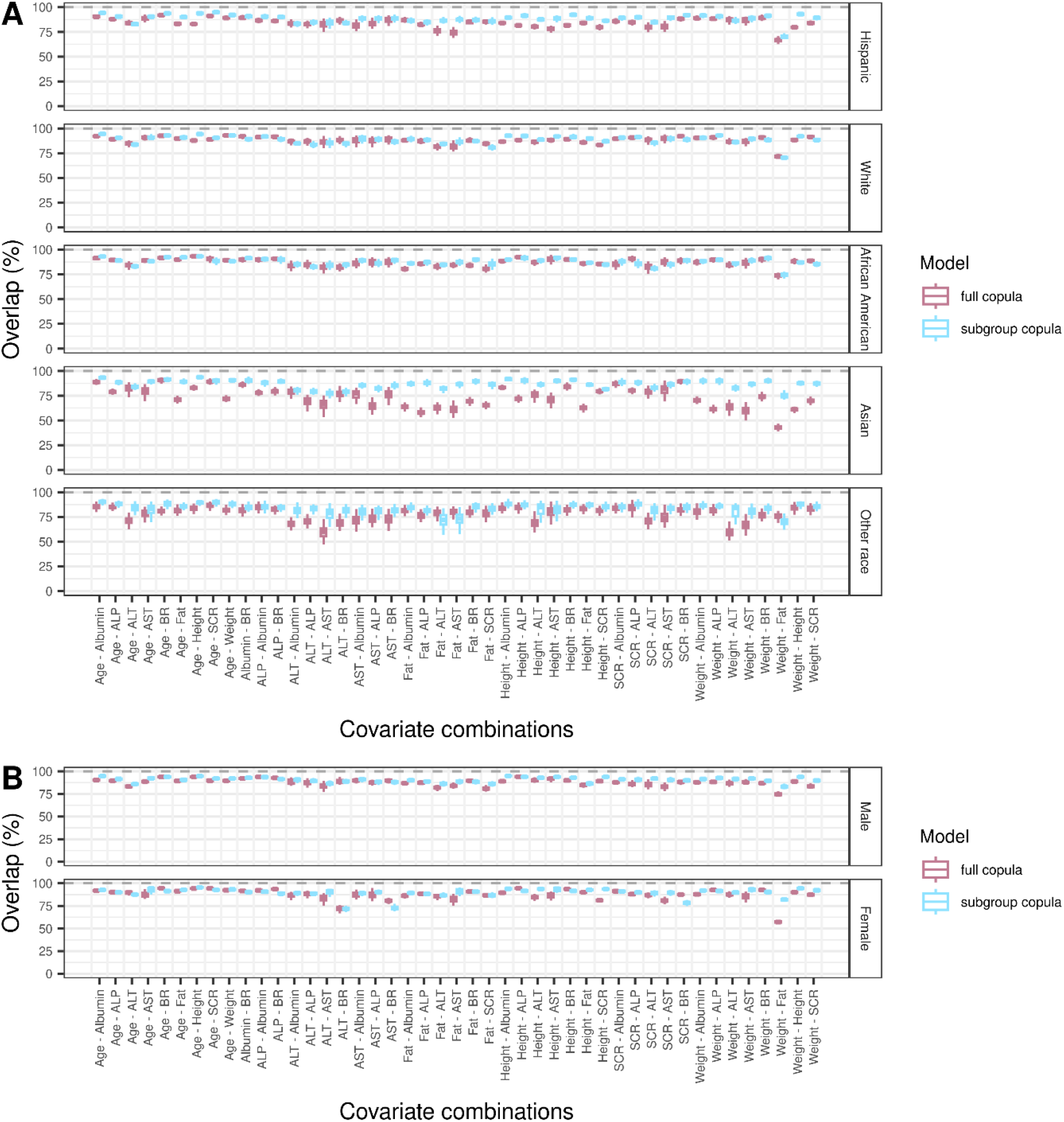
Overlap metric of each subgroup virtual population relative to corresponding real-world population. **A**. Overlap metrics calculated for each race-ethnicity subgroup population. **B**. Overlap metrics calculated for each sex subgroup population. The full copula was created utilizing the whole set of data, while subgroup copulas were developed based on each subgroup of data. Subgroup virtual populations were simulated 100 times using full copula (pink boxes) and subgroup copulas (blue boxes) for each. Error bars indicated the standard deviation of 100 simulations. Gray dashed lines indicate 100% overlap percentage.

#### 3.3.2 Sex subgroup analysis

In general, compared with subgroup copulas, the full copula model could well capture the margins and dependency structures in male and female populations. For marginal metrics, median relative errors of full copula were within the range [-0.19, 0.09] and [- 0.28, 0.07] for male and female populations (**Figure S3**). For comparison, subgroup copulas for male and female yielded median relative errors of [-0.11, 0.11] and [-0.03, 0.08]. The median overlap metric of full copula was calculated to be 88.5% and 88.8% for male and female populations (**Figure 4B**), while subgroup copulas achieved 91.1% and 90.8% overlap percentages for the two populations.

### 3.4 R shiny application

The copula covariate simulator (CoCoSim) web application was developed based on the NHANES copula and made available online (https://cocosim.lacdr.leidenuniv.nl/, **Figure 5**). Using this application, VPs can be generated online following these steps: (1) define the population of interest by selecting race-ethnicity, sex, age, and body mass index (BMI); (2) select the covariates of interest. Secondary covariates, including BMI, lean body weight, and estimated glomerular filtration rate, can be calculated based on the covariates in NHANES dataset; (3) select the number of individuals for simulation; (4) generate the VP and download the data.

**Figure 5.**
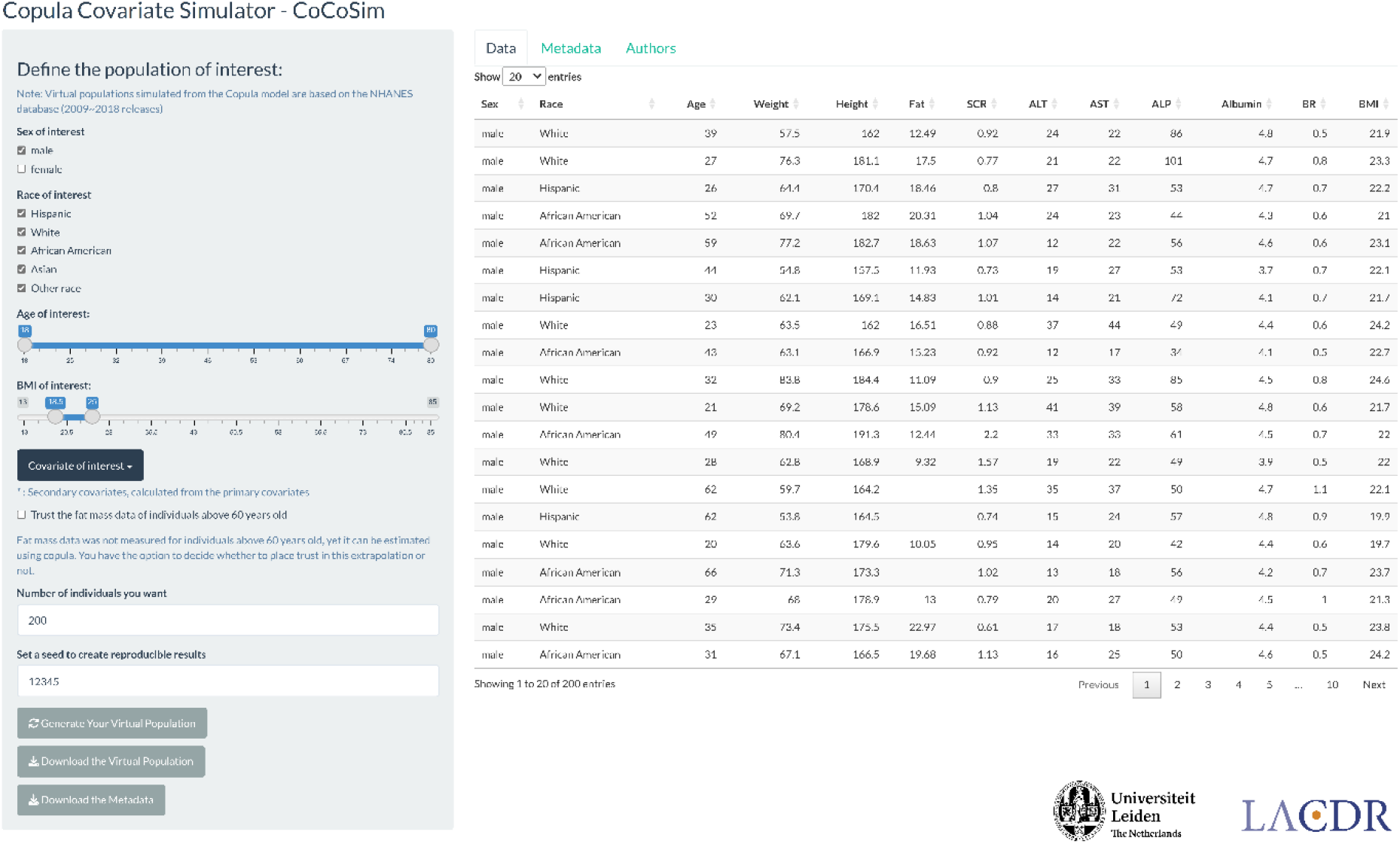
Interface of the R shiny application CoCoSim (https://cocosim.lacdr.leidenuniv.nl/). Virtual population can be generated according to user-defined characteristics based on NHANES copula (full copula).

With the app, users can generate virtual population with desired characteristics, including race-ethnicity, sex, age and BMI ranges. Generated virtual populations can then be used as covariate distributions for pharmacometric model-based simulations, such as for example as part of clinical trial simulations or dosing strategy optimization simulations.

## 4. Discussion

We developed a copula model for an adult population which adequately captured the covariate distributions as present in the NHANES database.

The tree structure of the NHANES copula revealed associations between commonly used covariates in population pharmacokinetics studies, which may help in the process of covariate model development. Identified associations were in line with the literature in which sex was found to influence height, weight, serum creatinine, and liver function biomarkers (total bilirubin and ALT) [20]. The correlation between covariates may explain the situations where sex may not be relevant as a covariate when the other covariates are included, since different PK or PD outcomes depend on underlying covariates (such as weight and serum creatinine) [21, 22].

To evaluate the performance of the developed copula model, we assessed whether the simulated population is realistic by comparing the marginal and dependency metrics between VPs and real-world population. Interestingly, we observed that the pair combinations of covariates that showed the largest errors of correlation differed from those showing the lowest overlap percentages. Pearson correlation quantifies linear association, while data sharing the same linear correlation could exhibit different dependency structures, and the overlap metric takes the shape or pattern of the dependency into account. The novelty of overlap metric lies in its first application of Jaccard index, a similarity measure between two data samples [23], to two-dimensional densities. Pearson correlation and overlap metric collectively depicted the joint behavior at a pairwise level and addressed different perspectives, and as such should be evaluated together when assessing copulas or investigating the similarity between two population.

In this study, we incorporated not only continuous but also categorical variables in the estimation of the NHANES copula. Currently, copula models for unordered categorical variables are not fully identifiable [17]. To include race-ethnicity (an unordered categorical variable) in copula, we estimated vine copulas by iterating through all possible orders of race-ethnicity and selected the model with the lowest AIC value. Since there were five categories in race-ethnicity, we considered 120 unique order possibilities of race-ethnicity categories, which was time-consuming and computationally expensive. Since this type of variable is common in clinical studies, such as disease classification, an algorithm that could efficiently deal with unordered categorical covariates is yet to be developed.

Copula models can be useful to support model-based dosing optimization or clinical trial simulation. For such applications, a focus on subjects with specific covariate characteristics usually exists [24, 25]. To this end, it is important to confirm whether a copula model correctly reflects covariate distributions for relevant population subgroups of interests. In our analysis, compared with subgroup copulas, the full copula model showed comparable performance across different race-ethnicity and sex subgroups except for Asian and Other race subgroups, likely due to the relatively small number of individuals within the entire dataset. The ability to adequately simulate subgroups from a large copula is of great importance since creating copulas for each subpopulation of interest, including e.g. different age and BMI ranges creates a nearly infinite amount of possible subgroups.

The NHANES population represents the non-institutionalized population of America and cannot be classified as healthy subjects or patients, indicating that the virtual population simulated from full copula should be interpreted with care. In this dataset, a significant portion of fat mass data was missing due to the age-eligible criterion (< 60 years old) of examination. However, copulas allowed for interpolation and extrapolation of VPs, as it supports the imputation of missing data via conditional density functions [26]. Of note, we removed the extrapolated fat mass data during the evaluation of copula performance. Although no significant bias was revealed in imputation analysis, simulated fat mass for people above 60 years old should be used with caution.

To make the full copula more accessible to the community, a web application was developed to facilitate the simulation of VPs with user-defined properties. This work served as a basis for building a copula library for sharing the copulas of various patient populations, such as obese, pregnant, or renally impaired patients, supporting simulation studies. Collaborative efforts could be initiated to gather large-scale data to build copulas for various target populations.

## 5. Conclusion

In this study, we demonstrated the development and evaluation of a copula model using NHANES database to simulate commonly used covariates in pharmacometric modeling, which can be used as part of clinical trial design and dose strategies optimization. A user-friendly web application was developed to facilitate the use of the developed copula model for covariate simulation.

## Data Availability

Code produced in this study are available online at https://github.com/vanhasseltlab/NHANES_copula.

## Authors’ Contributions

Conceptualization: J. G. Coen van Hasselt, Laura B. Zwep, Tingjie Guo; Methodology: Laura B. Zwep, Yuchen Guo; Formal analysis and investigation: Yuchen Guo; Writing - original draft preparation: Yuchen Guo; Writing - review and editing: J. G. Coen van Hasselt, Laura B. Zwep, Tingjie Guo, Catherijne A.J. Knibbe; Funding acquisition: J. G. Coen van Hasselt, Yuchen Guo; Supervision: J. G. Coen van Hasselt, Laura B. Zwep, Tingjie Guo, Catherijne A.J. Knibbe

