## Supplementary materials for "Generation of realistic virtual adult populations using a model-based copula approach"

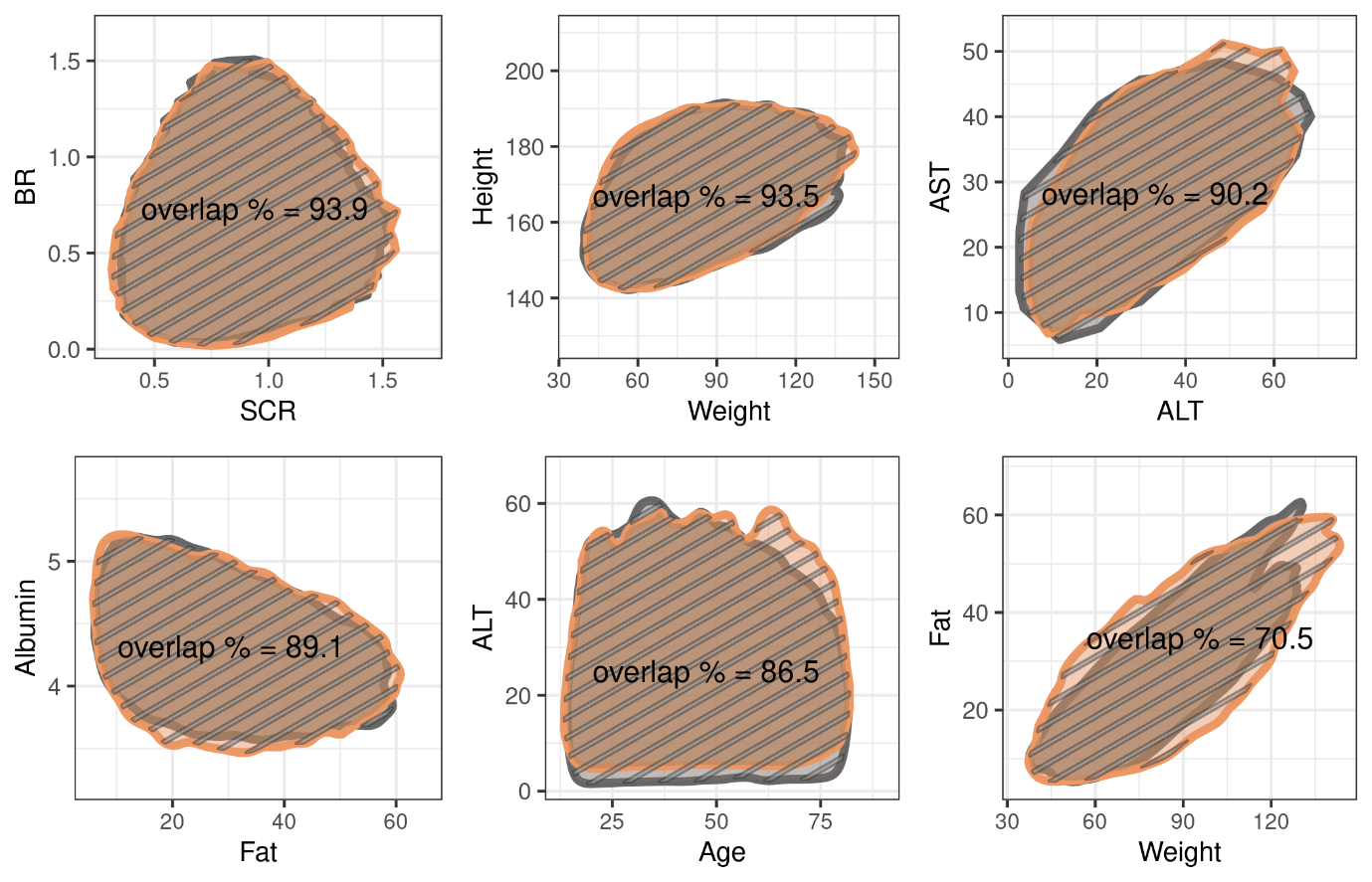
***Figure S1.*** *Illustration of overlap metric of 95^th^ contours in the selected covariate pairs. Orange and gray regions represent the areas of the 95^th^ contour of the observation population and virtual population. Brown and stripped area represents the overlap and union of these contours. Overlap percentage is calculated as the ratio of the intersection (brown area) over the union (stripped area).*

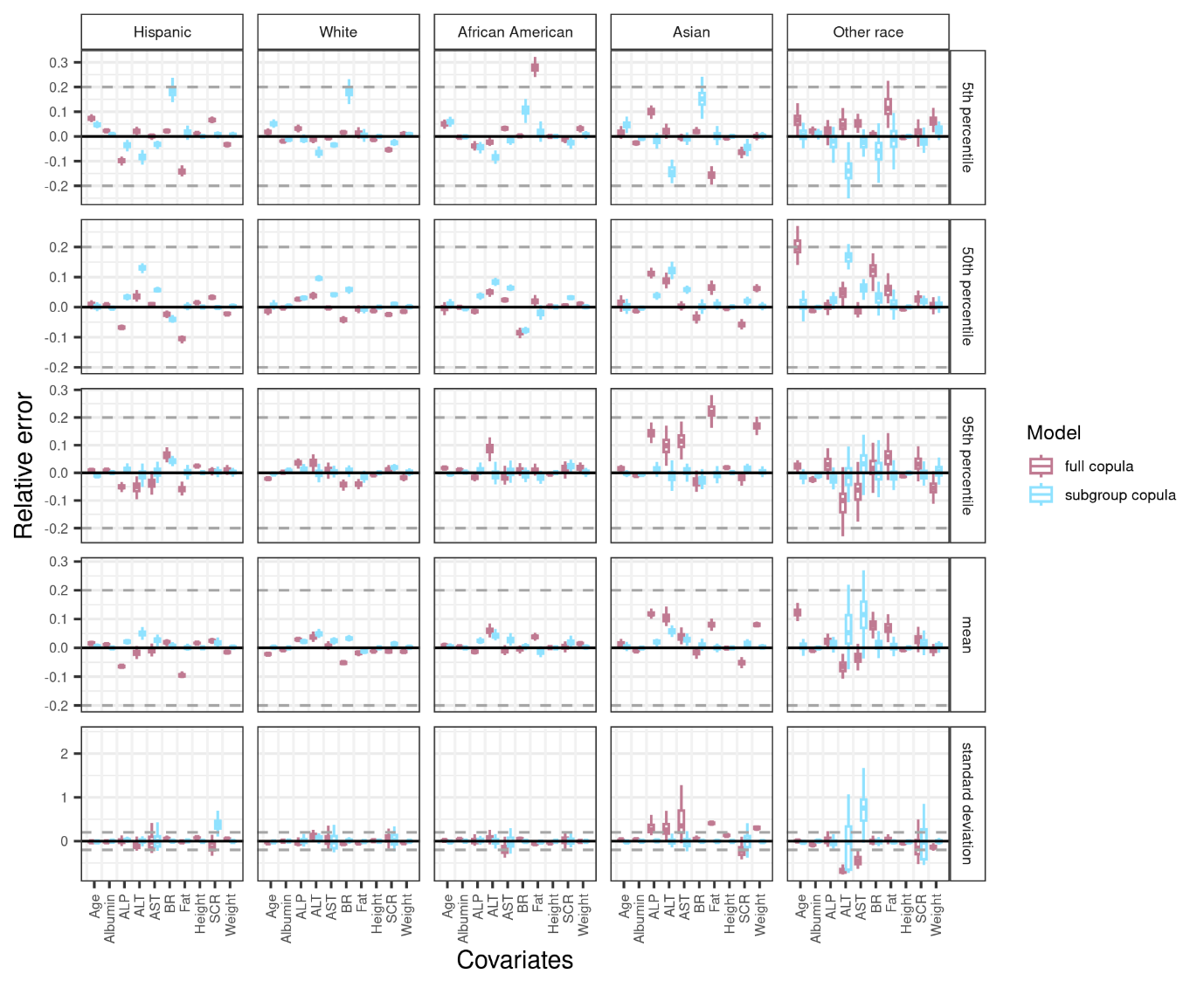

***Figure S2.*** *Relative error of marginal metrics in each race-ethnicity virtual subgroup population as compared to the statistics of corresponding real-world population. The full copula was created utilizing the whole set of data, while subgroup copulas were developed based on each subgroup of data. Virtual subgroup populations were simulated 100 times using full copula (pink boxes) and subgroup copulas (blue boxes) for each. Error bars indicated the standard deviation of 100 simulations. Gray dashed lines indicate ±20% relative error.*

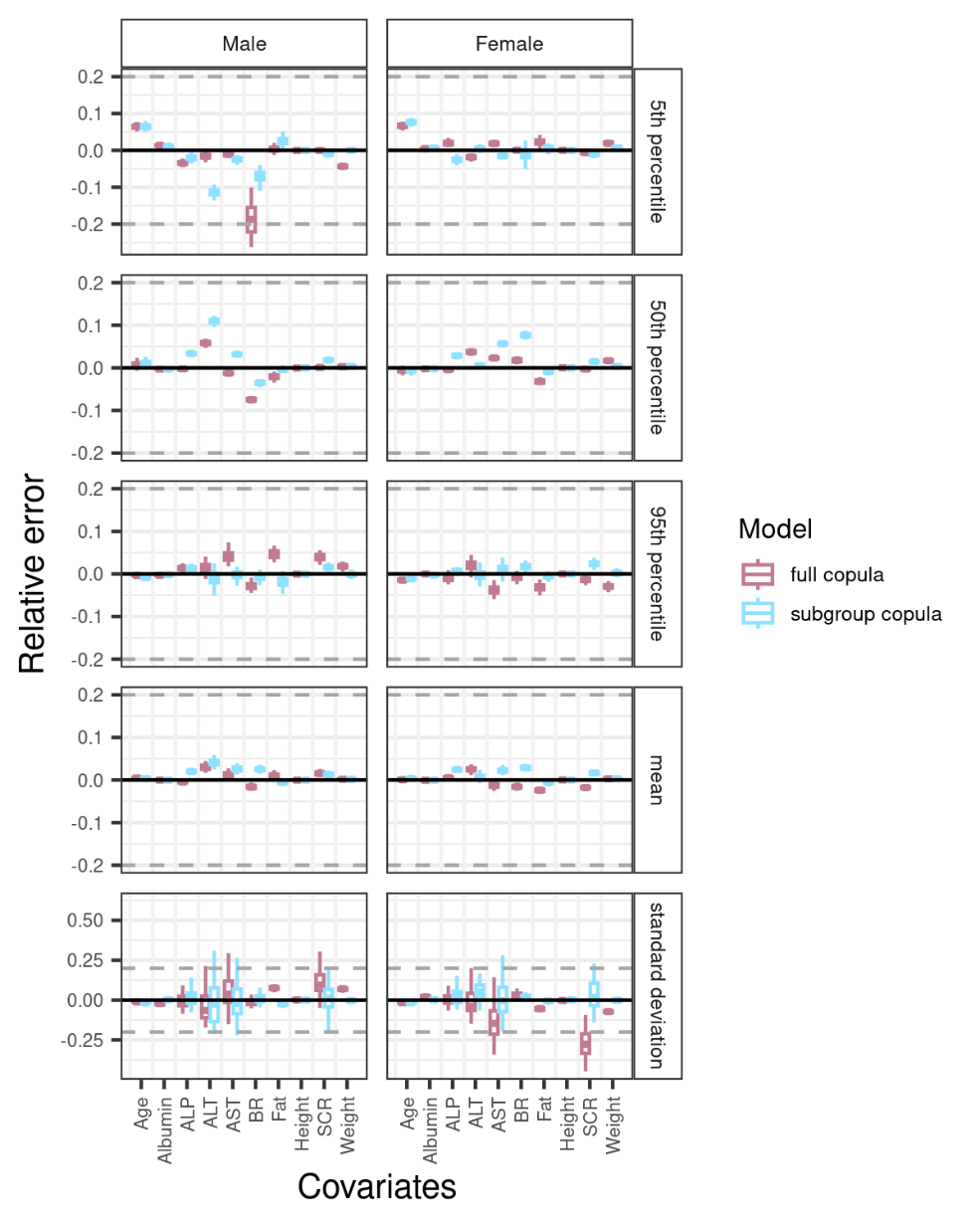

***Figure S3.*** *Relative error of marginal metrics in each sex virtual subgroup population as compared to the statistics of corresponding real-world population. Full copula was created utilizing the whole set of data, while subgroup copulas were developed based on each subgroup of data.* *Virtual subgroup populations were simulated 100 times using full copula (pink boxes) and subgroup copulas (blue boxes) for each. Error bars indicated the standard deviation of 100 simulations. Gray dashed lines indicate ±20% relative error.*

**Imputation analysis regarding missing fat mass data**

**Method**

In our real-world dataset, a significant portion of fat mass data was found to be missing due to the age-eligible criterion (< 60 years old) during the dual-energy x-ray absorptiometry examination. However, copulas allowed us to obtain a complete dataset for simulated VPs as it leveraged conditional density functions.

To provide insight into the reliability of simulated fat data for people above 60 years old, we applied the same missing data mechanism on the real-world dataset and investigated the performance of imputation of fat mass data for people with unmeasured fat mass data. We first extracted a complete dataset (N=11179) from the NHANES dataset. Based on this dataset, fat mass data for individuals aged between specific cut-off ages (47, 38, and 34) and 60 years old were removed to generate incomplete datasets with 30%, 50%, and 60% missing fat mass data.

Four vine copulas were developed based on the complete and incomplete datasets. Through 100 simulations, we assessed the extrapolation ability in generating the fat data for individuals lacking fat observation. For each level of missing fat data, we compared the marginal metrics of the fat data for people aged between cut-off ages and 60 years old in the real-world population, VPs simulated from complete-data-based copula, and VPs simulated from incomplete-data-based copula.

**Result**

Compared with observed fat mass data, fat mass simulated from the complete-data-based and incomplete-data-based copula models had most of marginal metrics within ±0.20 relative error (**Figure S4**), demonstrating the reliability of simulated fat mass data and the extrapolation ability of the copula model. With the increase of missing proportion (30%, 50% and 60%), incomplete-data-based simulations showed a tendency of underestimation, with –0.05, –0.09 and –0.13 median relative errors for 5^th^ percentile of fat mass, respectively; 50^th^ percentile (0.03, 0.00 and –0.03) and mean (0.03, 0.00 and –0.03) exhibited a similar trend.

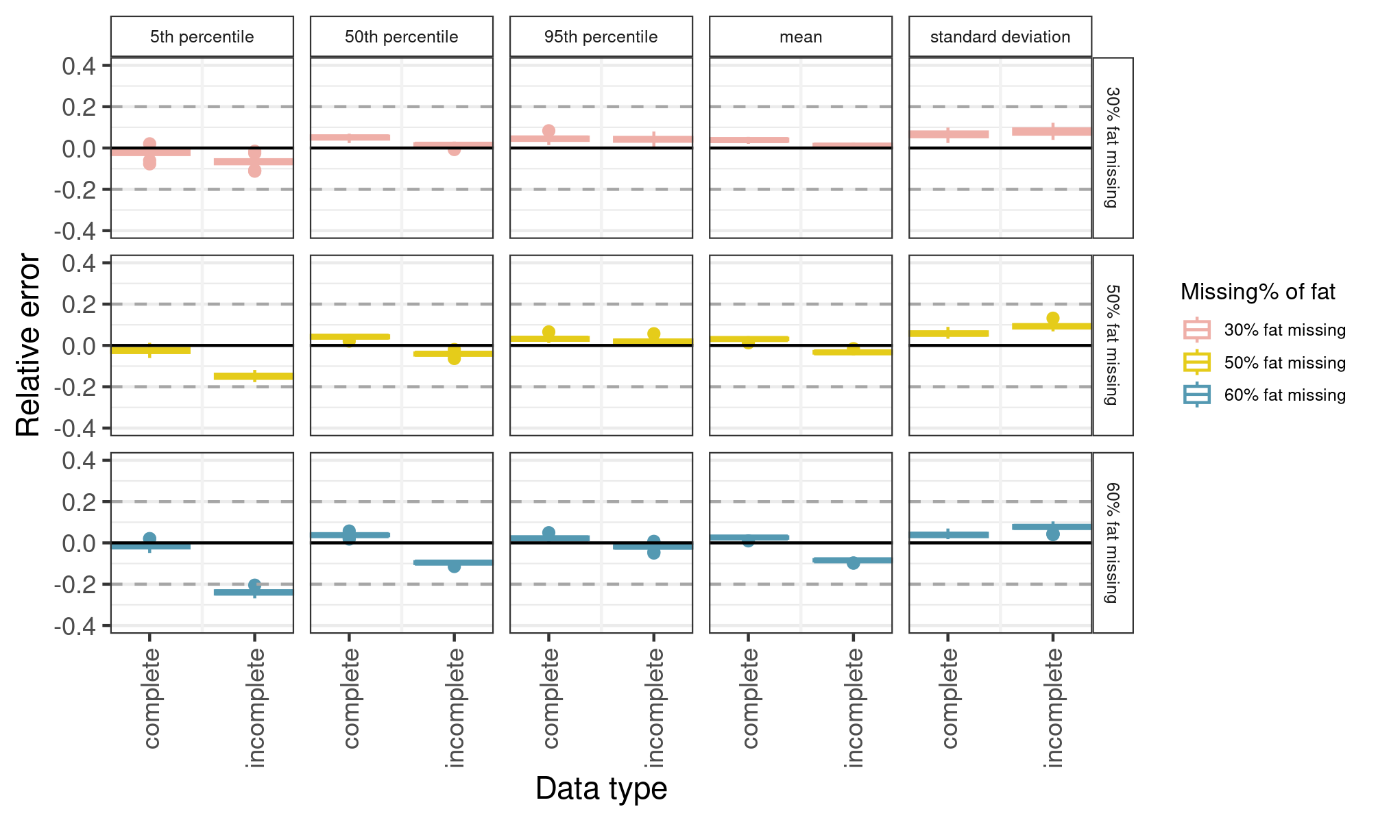

***Figure S4.*** *Relative error of marginal metrics of simulated fat mass data as compared to the statistics of observed fat mass data. Based on the complete dataset extracted from NHANES dataset, fat mass data for individuals aged between specific cut-off ages and 60 years old were removed to generate incomplete datasets with 30%, 50%, and 60% missing fat mass data. Each row represents the marginal metrics of fat data for people aged between a certain cut-off age and 60 years. Each subplot compared the performance of complete-data-based copula with incomplete-data-based copula. 30%, 50%, and 60% missing fat mass data corresponded to the cut-off ages of 47, 38, and 34 years old.* *Virtual population was simulated 100 times. Error bars indicated the standard deviation of 100 simulations.*
